# Trial protocol: A hybrid effectiveness-implementation study of Paediatric Autism Communication Therapy (PACT) in the Brazilian public health system

**DOI:** 10.64898/2026.06.18.26355880

**Authors:** Priscilla Brandi Gomes Godoy, Isabela Cóstola Windlin, Lais Menezes dos Santos Cardoso, Jennie Yoshida, Deborah Cristina Pereira Lopes, Karina dos Santos Arruda, Paulo Cesar Pinheiro de Oliveira, Isaac Figueira de Aquino, Raíssa Pazetto Alves, Thomas Nader Constancio, Ingrid Michelotti Aguiar, Thiago Figueiredo dos Santos, Daniela Lucas Nogueira Diniz, Larissa Andréa Lopes Cerqueira Lilge, Mariana Pereira de Castro, Jessica de Brito Sardinha Campos Proença, Guilherme de Almeida Prazeres, Daniela Molina-Avejonas, Erica Salomone, Kathy Leadbitter, Jonathan Green, Elizabeth Shephard

## Abstract

In Brazil, autistic children and their families face significant challenges in accessing evidence-based interventions, especially via the national public health service. Here, we report the protocol for a hybrid effectiveness-implementation trial, which will investigate the large-scale implementation of Paediatric Autism Communication Therapy (PACT) as well as its real-world effectiveness in Brazilian public health services for autistic children. Eligible professionals from public health services across the five regions of Brazil will be trained in PACT. They will deliver the intervention, which consists of 14 sessions delivered over 6 months, to dyads of autistic children and their caregivers in their regular clinical practice for the trial period of 18 months. We will use elements of three implementation science frameworks to systematically study factors that influence the implementation of PACT in this setting. To study effectiveness dyads will be randomised to receive PACT immediately or following a 6-month waitlist, stratified by healthcare service. Effectiveness outcomes (parent-child interaction, child adaptive social communication skills, caregiver well-being) will be collected pre- and post-PACT/waitlist and compared between groups. This trial will provide the first evidence on the effectiveness of PACT implemented in a public health system and the barriers and facilitators to such large-scale implementation.

**Lay abstract:** In Brazil, many autistic children and their families struggle to access early, evidence-based support through the public health system. This study aims to help change that. We will test how a well-established intervention, called Paediatric Autism Communication Therapy (PACT), can be delivered at a large scale in Brazilian public health services, and whether it works effectively in real-world conditions. PACT works by helping caregivers understand their autistic child’s communication and to respond in supportive ways that promote their child’s social communication development. For this project, professionals from public health services across all five geographical regions of Brazil will receive training to deliver PACT to autistic children and their caregivers as part of their routine clinical work. Each family will be offered 14 sessions over six months, and the study will run for a total of 18 months. To understand how PACT can be used successfully across the Brazilian public health services, we will examine what helps or hinders implementation using established frameworks from implementation science. We will also evaluate how well the intervention works by comparing families who start PACT immediately with those who begin after a short wait. We will assess changes in caregiver-child interactions, children’s social communication and well-being, and caregiver well-being. This will be the first study to examine an evidence-based therapy for autistic children and their caregivers implemented at scale in the Brazilian public health system. The findings will guide policymakers, health services, and communities on how to improve access to high-quality early support for autistic children and their families in Brazil.

## Introduction

Clinical guidelines emphasise the importance of early therapeutic support for autistic children (National Institute for Health and Care Excellence, 2013; 2021; Hyman, Levy, & Myers, 2020). While numerous early intervention models have been developed to support the development of autistic children (Sandbank et al., 2020) several characteristics are highlighted as essential. These include: robust evidence of efficacy from high-quality randomised trials with low risk of bias (Bottema-Beutel, 2023), appropriate implementation processes in community health services (Vivanti et al., 2018), supportive of neurodiversity and the priorities reported by autistic individuals and their families, which include mental health and well-being, community support and communication skills (Benevides et al., 2020; Dey et al., 2023; Flenik et al., 2023; Pellicano et al., 2014), generalisation of gains to different contexts of the child’s life (Bottema-Beutel et al., 2021), and family involvement in the therapeutic process (Gulsrud et al., 2016).

While there are intervention models that meet these criteria (Green, Leadbitter, Ainsworth et al., 2022), few are widely available to families via public health systems, especially in low-and-middle-income countries. In Brazil, for example, autistic children can receive free support through the Unified Health System (SUS) where clinical practice follows guidelines published by the Brazilian Ministry of Health concerning autism interventions; these guidelines highlight the importance of evidence-based interventions that support family involvement, respect neurodiversity and can be delivered via telehealth (Ministério da Saúde, 2023). However, it is well documented that autistic children spend months or years on SUS waiting lists before receiving assessment and support (dos Santos & Lima, 2024; Garcia et al., 2015; Paula et al., 2020) and families report a lack of specialized autism services and difficulty traveling to healthcare centers, which negatively impact family quality of life and financial health (dos Santos & Lima, 2024; Gomes et al., 2015; Lima et al., 2014; Paula et al., 2020). SUS healthcare professionals also report significant difficulties in meeting the demand for services due to being overworked, insufficient staffing, inadequate training and non-specialist service options (Bonfim et al., 2023; Faulin et al., 2021; Herr et al., 2024).

Thus, there remains a disconnect between the guiding principles of public policies aimed at the care of autistic children and their families in Brazil and the actual practices. We propose that the implementation of Paediatric Autism Communication Therapy (PACT, Green et al., 2010) could improve the provision of support for autistic children and their families in the Brazilian SUS. PACT is a caregiver-mediated therapy designed to improve the sociocommunicative and broader development of autistic children aged 2 to 11 years (Pickles et al., 2016). The goal is to support development by increasing caregiver sensitivity and responsiveness to the child’s individual communication and interaction style and their awareness of wider autistic development. It will involve 14 fortnightly sessions with the caregiver, during which a trained therapist uses video-feedback of naturalistic interactions between the caregiver-child dyad to support the caregiver to increase their sensitivity to the child’s social communication skills and adapt their communication style to that of the child. Caregivers will undertake home practice of the PACT strategies to generalize the therapy gains.

As a caregiver-mediated intervention, PACT requires less time from healthcare professionals than many other models, whilst providing strong evidence of long-term sustained effectiveness (Pickles et al., 2016). PACT can be delivered online (Geoffray et al., 2025; Green, Leadbitter, Ellis et al., 2022), meaning it can reach families living in remote areas. Evidence for long-term effectiveness comes from formal RCT (randomized controlled trial) follow up (Pickles et al 2016), as well as effects on child social communication development, caregiver-child interaction and family well-being (Green, Leadbitter, Ellis et al., 2022; Leadbitter et al 2018). PACT has also demonstrated good cost-effectiveness into middle childhood on economic modelling (Tinelli et al 2023) and has been successfully implemented in public health services in Europe and, after cultural adaptation, in South Asia (Balabanovska et al., 2025; Divan et al., 2015, 2019; Rahman et al., 2016).

Considering these characteristics of PACT, we propose that this intervention provides a viable, economical, and effective therapeutic option for autistic children and their families within the Brazilian public health system. Recently, we conducted a small-scale qualitative implementation study of PACT in Brazil, in which we identified barriers to its implementation in this context and made cultural adaptations to the PACT protocol (Godoy et al., 2024). Main adaptations involved adjusting the frequency of PACT sessions to maximise caregiver engagement at the beginning of the therapy and the use of culturally appropriate language, illustrative examples of PACT strategies and toy recommendations for caregiver-child interactions (Godoy et al., 2024). We showed that this adapted version of PACT presented good acceptability and feasibility amongst families and healthcare professionals in the state of São Paulo (Godoy et al., 2024). The current trial will build on this work by (1) testing the real-world effectiveness of PACT in improving child social-communication skills, caregiver-child interaction and family well-being as implemented at national scale in the Brazilian SUS, and (2) use elements of three implementation frameworks to systematically investigate barriers and facilitators to the implementation of PACT in SUS.

## Methods

### Ethical approval and trial registration

Ethical approval for this trial was granted by the Research Ethics Committee of the Institute of Psychology, University of São Paulo (Ref: 86935425.6.0000.5561). Adult participants will provide written informed consent before completing any study measure. The study was prospectively registered in the international ISRCTN registry for clinical trials (ref: ISRCTN13161676). The trial protocol is available as a preprint in English with a supplementary version translated to Portuguese available on the Open Science Framework (Godoy & Shephard, 2025).

### Trial funding, management and monitoring

This trial is funded by a research grant from the São Paulo Research Foundation (ref 2025/03106-1). The funder has no role in the trial beyond financial support. The *Trial Sponsor* is the University of São Paulo; the principal investigator (ES) is responsible for the overall conduct and coordination of the trial. The *Trial Management Group* is composed of the senior researchers of the trial (ESh, PBGG, ESa, DAM, KL, JG) and will oversee day-to-day operations, ensure protocol adherence, and monitor recruitment, data collection, handling and analysis. The *Steering Committee* includes the investigators ESh and PBGG, three independent researchers with expertise in clinical trials and implementation science from the UK and Brazil and one member of the lay public from Brazil. The Steering Committee will meet every six months to assess trial progress, adherence to the trial protocol and analysis plan, assess participant safety, advise on decisions and actions and ensure transparent and timely reporting of trial findings. The *Data and Safety Monitoring Board* composed of two independent Brazilian researchers will review data on adverse events and fidelity of PACT delivery every four months to monitor participant safety, trial validity and data integrity.

In line with participatory research principles (Staniszewska et al., 2017) and recommendations for meaningful autistic involvement in research (Fletcher-Watson et al., 2019), the trial includes a *Community Advisory Board* (CAB) composed of 12 autistic adults and two parents of autistic youth resident in Brazil who co-designed the protocol for the trial. Members of the CAB will continue to advise on the conduct of the study and participate in data analysis via regular meetings and/or asynchronous communications with the Brazilian main researchers (ES and PBGG). In Brazil we have encountered significant challenges maintaining participatory involvement of autistic adults (Tafla et al., 2026). Therefore, we recruited a large number of autistic adults to participate in the CAB to ensure we will have continuous community representation throughout the trial, despite expected fluctuations in individual participation.

### Design

This is a type I hybrid effectiveness-implementation trial (Curran et al., 2012) to test the effectiveness of PACT in the Brazilian public health system (SUS) while collecting data on implementation barriers and facilitators. We will recruit SUS services that support autistic children across the five regions of Brazil (at least two services per region). All eligible professionals of the services will be trained to manual fidelity in PACT and deliver the intervention (14 sessions over 6 months per family) to dyads of autistic children aged 2-10 years and their caregivers in their regular clinical practice over an 18-month period. After consent, dyads will be randomised within each service to either receive PACT or enter a 6-month waitlist period during which they will receive care-as-usual (CAU). Primary and secondary outcomes will be assessed at pre-randomisation baseline (hereafter, T1) and immediately following the six months of intervention/waitlist (hereafter, T2). Data on barriers and facilitators to implementation will be collected throughout the 18-month trial period.

### Settings

PACT will be implemented in SUS services that provide support to autistic children distributed across the five geographical regions of Brazil (North, Northeast, Midwest, Southeast, South). PACT training for professionals will be conducted online. The delivery of PACT in each service will be in-clinic or online, following the individual preferences of the professionals and families. Data collection of effectiveness and implementation outcomes will be conducted online by psychologists based in São Paulo.

### Study population

Participants will be healthcare professionals and managers of SUS services and autistic children and their primary caregivers who are attended at these services. The SUS services will be recruited via institutional social media advertisements and word-of-mouth amongst healthcare professionals. Managers of interested services will complete an online questionnaire to assess the eligibility of the service to participate in the trial. Eligible services will be those that offer clinical support to autistic children delivered by psychologists, speech therapists and/or occupational therapists and those that represent sociodemographic and cultural diversity across the five regions of Brazil. Services will be excluded if they do not offer supports for autistic children, are temporary services or depend on research funding for intervention provision.

Within the SUS services, managers will participate in the assessments of implementation barriers and facilitators, as well as coordinate and monitor the implementation of PACT by healthcare professionals in the service. The healthcare professionals of the services will be eligible to receive training in PACT and deliver the intervention in their normal clinical practice if they are psychologists, speech and language therapists or occupational therapists who already work directly with autistic children within the service and hold a non-temporary contract. These professionals will also complete assessments of barriers and facilitators to implementation of PACT and liaise with the research team to facilitate the effectiveness aspect of the trial. Healthcare professionals will be excluded if they have no experience of working with autistic children or do not wish to work with PACT.

Autistic children attending the public health services will be considered eligible for inclusion if they: (1) hold a diagnosis of autism *or* are undergoing assessment for autism *and* score above the clinical threshold for autism on the Social Communication Questionnaire (SCQ, Rutter et al., 2003), which will be administered to the primary caregiver by the research team; (2) are aged 2 years 0 months to 10 years 11 months at the point of recruitment (the age-range for PACT); and (3) their primary caregiver agrees to participate in the trial. Caregiver-child dyads will be excluded from the study if: (1) they do not wish to receive PACT; (2) the primary caregiver does not speak Portuguese sufficiently fluently to be able to receive PACT; or (3) the primary caregiver has significant needs of their own that require urgent clinical attention, in which case the family will be invited to participate after these needs have been addressed. We will not exclude children with concurrent diagnoses of other conditions since our aim is to evaluate the real-world implementation and effectiveness of PACT.

### Implementation procedures

Online meetings will be conducted with managers of eligible services to explain the trial, the PACT intervention and discuss feasibility of the service’s participation. Eligible services that agree to participate will provide an authorisation letter. The service manager and eligible healthcare professionals will provide written informed consent. Next, healthcare professionals will be trained to manual fidelity in PACT by accredited Brazilian PACT trainers (ESh and PBGG) following standard PACT training procedures, which include a 4-hour asynchronous e-learning module, a practical course taught synchronously across four half-days, and post-course group and individual supervision of 1-2 clinical cases. Training takes approximately four months and will be delivered remotely in Portuguese. Healthcare professionals will not begin delivering PACT within their service until they complete the training and meet the criteria for delivery according to intervention manual fidelity as assessed using the PACT Fidelity Checklist (Aldred et al., 2024).

Once healthcare professionals are trained to fidelity, they will deliver PACT to eligible families for the implementation period of 18 months. Each service will adopt its own strategy for how they will deliver and offer PACT to families consistent with their existing practices, with support from the investigators ESh and PBGG. All families that meet the inclusion and exclusion criteria will be invited by the service healthcare professionals to participate in the study. Families who agree to participate will be contacted by the research team about the study. Written informed consent will be obtained from caregivers. Children’s assent to complete study measures and PACT session activities will be gauged continually based on their thoughts, words, actions and feelings. Each participating service will receive one hour of group supervision per month for therapists delivering PACT during the 18-month trial period. One PACT session per therapist per service will be randomly selected for fidelity assessment every three months using the standard PACT Fidelity Rating Scale.

### Randomization and blinding

After T1 assessments, each caregiver-child dyad will be randomly allocated to PACT or Waitlist+CAU by stratified randomisation using Variance Minimisation (Sella et al., 2021). Randomisation will be conducted in MATLAB (MathWorks, Natick, MA) by an independent researcher, not involved in other aspects of the trial, and will be stratified by site, i.e., the SUS service where the dyad is attended. The results of the randomisation will be passed to the implementation lead (PBGG) who will feed this back to health service, which will coordinate the local implementation of PACT and CAU with the family.

Researchers conducting the assessments will be blinded to allocation. Caregivers will be reminded by the trial manager that they must not reveal their allocation to the researchers. The primary outcome will be coded from video recordings by researchers blinded to intervention allocation. The secondary outcomes will be assessed by caregiver-report and are non-blinded, since families cannot be blinded to allocation.

### PACT Intervention

PACT will be delivered to families according to its protocol adapted for Brazil (Aldred et al., 2024; Godoy et al., 2024). This consists of 14 1/1.5-hour sessions between the therapist and the caregiver-child dyad delivered over six months, with weekly sessions for the first month followed by fortnightly sessions for the last five months. In each session, the therapist uses video-recorded naturalistic interactions between the caregiver and child to support the caregiver in recognising and understanding their child’s communication and implement strategies that support communication development. Families will also be asked to undertake daily home practice of the strategies for 30 minutes per day. Sessions will be structured according to the PACT intervention manual (Aldred et al., 2024). Further details on the PACT intervention are presented in the Supplementary Materials. Families assigned to PACT will continue to receive care-as-usual (CAU) whilst also receiving PACT. Details of care received as reported by caregivers and SUS service professionals will be registered by an unblinded researcher leading the implementation of PACT (PBGG) at T2 assessments.

### Waitlist + CAU

Families allocated to the Waitlist + CAU group will receive their usual care (CAU) from the SUS service and any other support they normally receive in the community or privately for six months, after which they will complete T2 assessments and then will be offered PACT by the SUS service. Effects of PACT in Waitlist + CAU families will not be studied in this trial. Details of care received as reported by caregivers and SUS service professionals will be registered by an unblinded researcher leading the implementation of PACT (PBGG) at T2 assessments.

### Harms

Participant safety and trial integrity will be monitored every four months by the Data and Safety Monitoring Board. Potential adverse events will be defined as increased emotional distress in the caregiver or child, increased caregiver stress, family conflict arising from changes in interactions or routines, or other unforeseen psychosocial reactions. Serious adverse events will be defined as any event in caregiver or child involving risk to life, hospitalisation, increased functional impairment, new/increased child protection concerns or serious data breaches. Adverse event monitoring will be through active surveillance (a brief post-session checklist completed by PACT therapists, presented in the Supplementary Materials) and passive surveillance (spontaneous caregiver reports to researchers or therapists and spontaneous therapist feedback to the researchers). Adverse events will be documented using a standard report form and handled using standardised procedures (see the Supplementary Materials). Implementation-related incidents (e.g., therapist overload, recording failures, logistic barriers) will be logged by the implementation lead (PBGG) as “implementation harms” and included in the 4-monthly safety and quality reports.

### Effectiveness outcome measures

#### Primary outcome

The primary effectiveness outcome is the synchrony of caregiver-child interaction quantified using the Dyadic Communication Measure for Autism (DCMA, Aldred et al., 2004) assessed at T1 and T2. The DCMA subscale *Caregiver Synchronous Communication Acts to the Child* (hereafter, *Synchrony*) will be coded from 10-minute video recordings of a free-play interaction between the caregiver and child conducted in the family’s home. Synchrony is defined as the proportion of verbal communication acts produced by the caregiver that are non-directive (comments, statements or acknowledgements), attuned to the child’s focus of attention, and which maintain the flow of the interaction (Aldred et al., 2004). Coding will be conducted by researchers trained to reliability against a master coder (expert in DCMA), blinded to allocation. Fortnightly consensus meetings between coders will be conducted to facilitate trouble-shooting and reduce drift in coding from the DCMA manual. 20% of videos will be double-coded to assess inter-coder reliability.

The choice of Synchrony coded from caregiver-child interactions in the family home as the primary outcome in this trial is not aligned with recommendations that primary outcomes of autism intervention trials should be distal to the intervention context and pose low risk of detection bias (LaPoint et al., 2025; Sandbank et al., 2023). However, we chose Synchrony as the primary outcome for three reasons: (1) caregiver-child interaction data can be collected remotely, which will be critical for the feasibility of data collection across a large number of families from public health services distributed throughout Brazil, including some remote and inaccessible locations; (2) outcome measures were selected in coproduction with autistic adults and parents of autistic children, who emphasised that the effectiveness of PACT should be assessed in terms of how well it strengthens family bonds and acceptance, as well as enhances the child’s meaningful communication and emotional well-being; (3) mechanistic analyses of previous PACT RCTs have shown that caregiver synchrony is the mediator of improvements in child communication development (Green, Leadbitter, Ellis et al., 2022; Pickles et al., 2016). To reduce confounding as much as possible, we will request that families use a specific set of toys for the DCMA interactions and that these are not used during PACT practice sessions.

### Secondary outcomes

#### Child communication initiations

The proportion of total communication acts that are child initiations will be coded at T1 and T2 using the DCMA (Aldred et al., 2004) from video-recordings of the naturalistic caregiver-child interactions as described above for the primary outcome. Child Initiations are defined as verbal or non-verbal communication acts by the child which start an interaction with the caregiver. This measure was shown to be important in mechanistic analyses of PACT: caregiver synchrony increased child initiations, which in turn increased social communication skills beyond the context of the caregiver-child dyad (Green, Leadbitter, Ellis et al., 2022; Pickles et al., 2016). Coding will follow the same procedures as described above for the primary outcome.

#### Child adaptive functioning

Children’s adaptive behaviour will be assessed at T1 and T2 using the Vineland Adaptive Behavior Scales 3rd Edition (Vineland-3, Sparrow et al., 2016) Standard Scores for the domains of Communication, Socialization and Daily Living and the Adaptive Behavior Composite. The Vineland 3 will be administered in interview format with the primary caregiver by researchers blinded to allocation.

#### Child well-being

Children’s well-being will be assessed by caregiver-report at T1 and T2 using the Brazilian Portuguese version of the Strengths and Difficulties Questionnaire (SDQ, Goodman, 1997) P2-4 version for children aged 2-3 years and P4-17 Version for children aged 4-10 years. Total scores for the Emotion, Conduct, Hyperactivity/Attention, Peer Relationship and Prosocial scales will be used in analyses. The SDQ will be administered in interview format with the primary caregiver by researchers blinded to allocation.

#### Caregiver well-being

Caregiver well-being will be assessed at T1 and T2 using the Portuguese version of the World Health Organisation-Five Well-Being Index (WHO, 2024). The total score will be used in analyses. The scale will be administered in interview format with the primary caregiver by researchers blinded to allocation.

#### Family well-being

Family well-being will be assessed at T1 and T2 using the Autism Family Experience Questionnaire (AFEQ, Leadbitter et al., 2018), which has been used in previous PACT trials and translated to Brazilian Portuguese (Faker, de Paula & Tostes, 2024). The AFEQ will be administered to the primary caregiver by researchers blinded to allocation.

#### Caregiver reflective functioning

Caregiver capacity to reflect on and understand their own and their child’s internal mental experiences and how they relate to their child’s behaviour will be measured at T1 and T2 using the Portuguese version of the Parental Reflective Functioning Questionnaire (PRFQ, Luyten et al., 2017; Moreira & Fonseca, 2023). Scores for the three PFRQ subscales, Pre-mentalising Modes of Mental States, Certainty About Mental States, and Curiosity in Mental States, will be used in analyses. The PFRQ will be administered to the primary caregiver in interview format by researchers blinded to allocation.

#### Child functioning at school

To explore children’s functioning in the educational environment, qualitative data will be obtained from school-based educational planning documents used in Brazilian inclusive education: the Plano Educacional Individualizado (PEI – Individualized Educational Plan) and the Plano de Atendimento Educacional Especializado (PAEE – Specialized Educational Support Plan). In Brazil, these documents are commonly developed within the framework of inclusive education policies and are used by schools to plan and monitor individualized educational strategies for students with disabilities, including autistic children.

The PEI is typically developed collaboratively by classroom teachers, special education professionals, and sometimes the family, and outlines individualized learning goals, accommodations, pedagogical strategies, and adaptations required to support the student’s participation in the general education curriculum. The PAEE is a complementary document used within the Atendimento Educacional Especializado (Specialized Educational Support Services), a component of the Brazilian inclusive education system that provides additional educational support to students with disabilities, usually delivered by specialized teachers in resource rooms or support centers within schools. The PAEE usually includes information about the student’s learning profile, communication and social interaction patterns, observed barriers to participation in the classroom, and the educational strategies implemented to support development.

Although the use of these documents is recommended by Brazilian educational policies supporting inclusive education, their implementation across schools is heterogeneous, reflecting differences in local resources, teacher training, and institutional practices. When available, these documents provide valuable naturalistic descriptions of the child’s functioning in the school environment from the perspective of educational professionals.

For the purposes of this study, parents will be invited to request copies of their child’s PEI and/or PAEE from the school using a standardized letter prepared by the research team. The documents will be anonymised and analysed qualitatively using thematic analysis to explore teachers’ descriptions of the child’s socio-communicative behaviours, participation in classroom activities, interaction with peers and teachers, and perceived educational needs. This analysis will provide an ecological perspective on child functioning in the school context, complementing the clinical and family-reported outcomes collected in the study.

### Implementation outcome measures

We will use elements of three implementation science frameworks to investigate the implementation of PACT in SUS. Specifically, we will use outcomes aligned with selected domains of the *Consolidated Framework for Implementation Research* (CFIR; Damschroder et al., 2022) to study *contextual* barriers and facilitators to implementing PACT in SUS (see Table 1). We will use outcomes in line with selected domains of the *Theoretical Domains Framework* (TDF; Cane et al., 2012) to investigate barriers and facilitators related to *behavioural changes* (Table 1). Finally, we will use the broad implementation outcome measures from the *Implementation Outcomes Framework* (IOF; Proctor et al., 2011) to assess the *success of implementing* PACT in SUS (Table 1). By selecting specific elements of these frameworks and combining them we will obtain a multi-level understanding of how organisational, individual, and system factors interact to shape the success and sustainability of implementing PACT in SUS, capturing the full pathway from *context* to *mechanism* to *outcome* (Birken et al., 2017). Qualitative and quantitative data on these implementation outcomes will be collected using semi-structured interviews and Likert ratings, as described in the following sections.

**Table 1.**
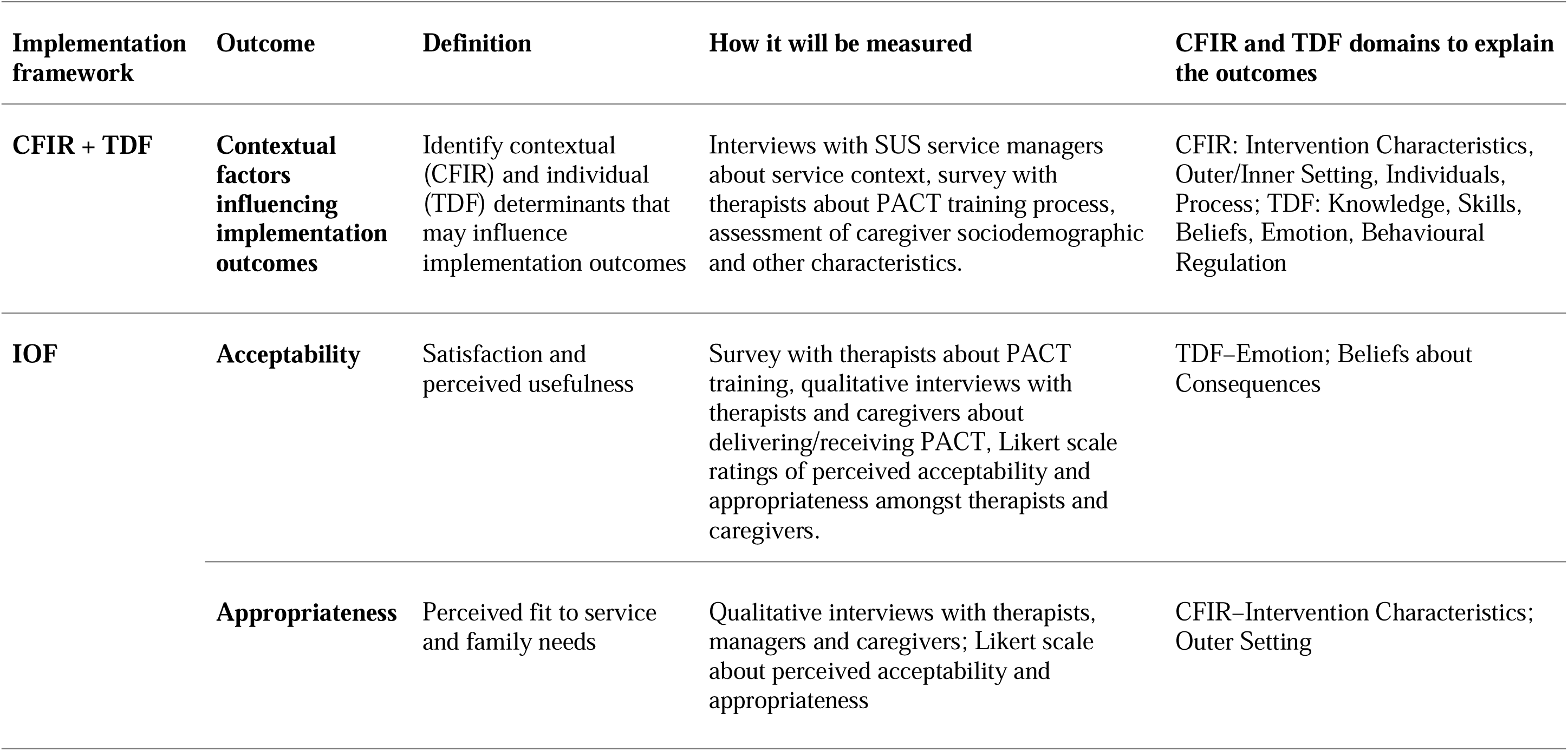
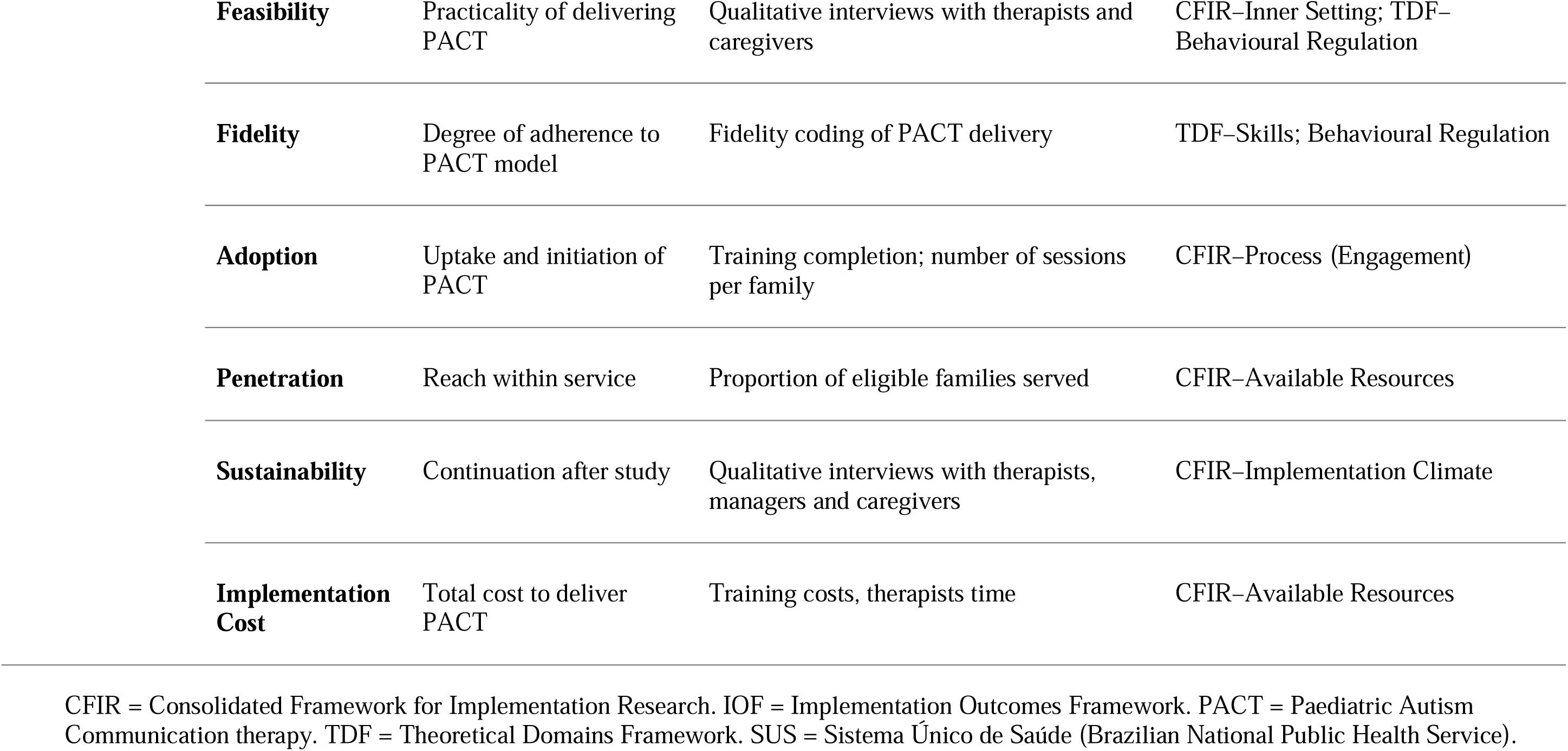
Implementation outcomes, frameworks, definitions, domains assessed, and data collection methods.

#### Implementation interviews with SUS service managers

Before implementation begins, interviews will be conducted with managers/coordinators of each SUS service. The interviews will be guided by CFIR domains to identify contextual determinants that influence IOF outcomes, including leadership engagement, resource availability, organisational culture, workflow constraints, compatibility of PACT with existing service structures, and anticipated barriers and facilitators to adoption. These contextual data will facilitate interpretation of implementation outcomes across sites. Interviews are expected to last 30-40 minutes and will follow the CFIR-based guide in the Supplementary Materials.

#### Implementation interviews with therapists and caregivers

At the end of the implementation period, the SUS therapists and caregivers will participate in semi-structured interviews about their experiences of delivering or receiving PACT, respectively. The interviews will be organised around TDF and IOF domains to assess behavioural mechanisms (TDF) that influence implementation (see Table 2) and the core IOF implementation outcomes listed in Table 1. Interviews are expected to last 30-45 minutes and will follow the guides in the Supplementary Materials.

**Table 2.**
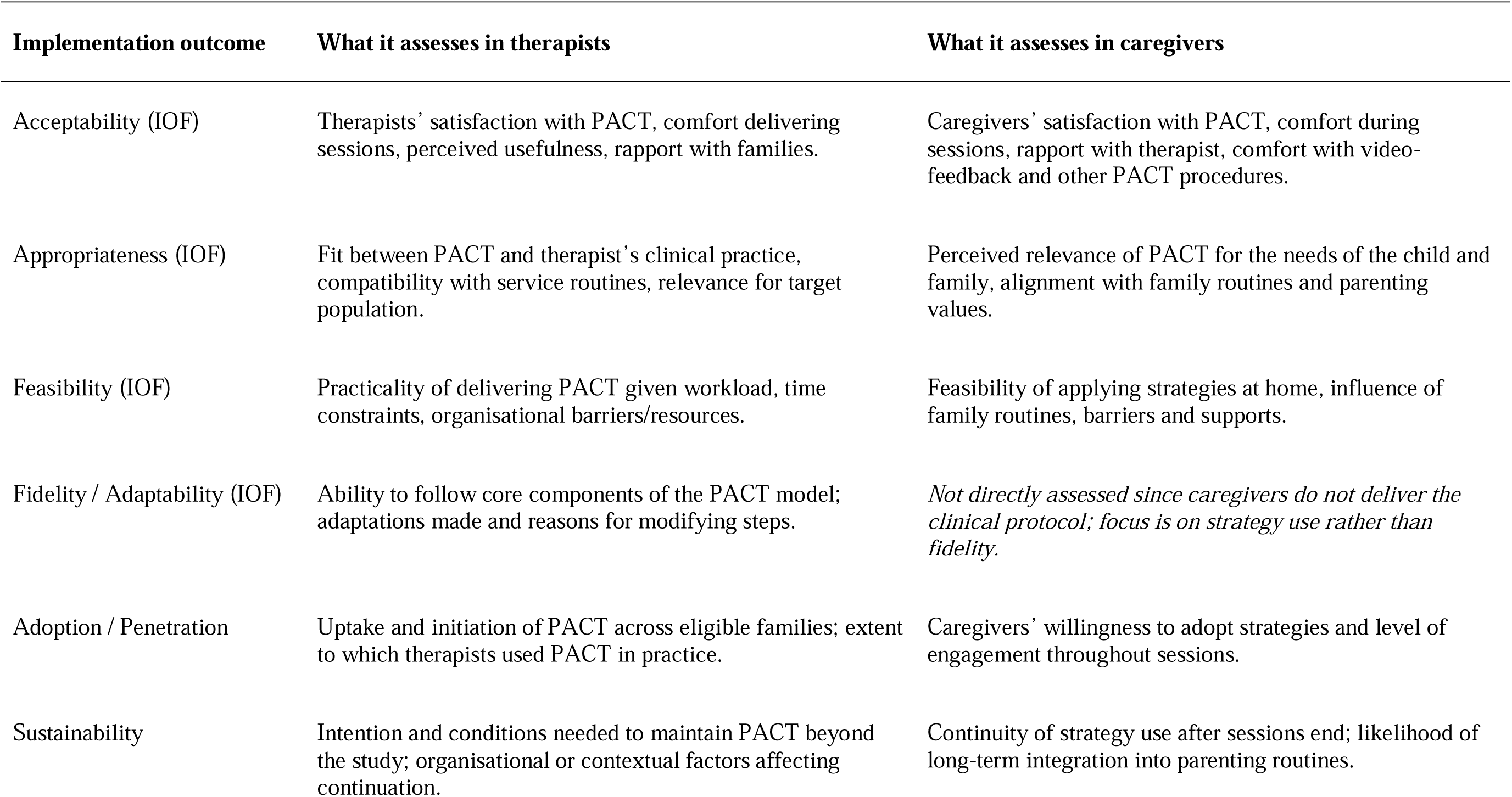
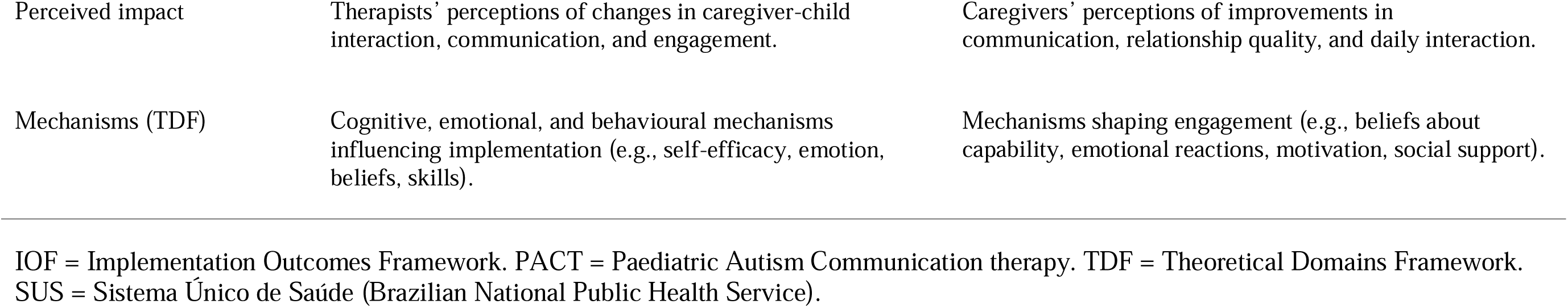
Summary of what implementation outcomes will measure in SUS therapists and caregivers.

| Implementation outcome | What it assesses in therapists | What it assesses in caregivers |
| --- | --- | --- |
| Acceptability (IOF) | Therapists' satisfaction with PACT, comfort delivering sessions, perceived usefulness, rapport with families. | Caregivers' satisfaction with PACT, comfort during sessions, rapport with therapist, comfort with video-feedback and other PACT procedures. |
| Appropriateness (IOF) | Fit between PACT and therapist's clinical practice, compatibility with service routines, relevance for target population. | Perceived relevance of PACT for the needs of the child and family, alignment with family routines and parenting values. |
| Feasibility (IOF) | Practicality of delivering PACT given workload, time constraints, organisational barriers/resources. | Feasibility of applying strategies at home, influence of family routines, barriers and supports. |
| Fidelity / Adaptability (IOF) | Ability to follow core components of the PACT model; adaptations made and reasons for modifying steps. | *Not directly assessed since caregivers do not deliver the clinical protocol; focus is on strategy use rather than fidelity.* |
| Adoption / Penetration | Uptake and initiation of PACT across eligible families; extent to which therapists used PACT in practice. | Caregivers' willingness to adopt strategies and level of engagement throughout sessions. |
| Sustainability | Intention and conditions needed to maintain PACT beyond the study; organisational or contextual factors affecting continuation. | Continuity of strategy use after sessions end; likelihood of long-term integration into parenting routines. |
| Perceived impact | Therapists' perceptions of changes in caregiver-child interaction, communication, and engagement. | Caregivers' perceptions of improvements in communication, relationship quality, and daily interaction. |
| Mechanisms (TDF) | Cognitive, emotional, and behavioural mechanisms influencing implementation (e.g., self-efficacy, emotion, beliefs, skills). | Mechanisms shaping engagement (e.g., beliefs about capability, emotional reactions, motivation, social support). |

IOF = Implementation Outcomes Framework. PACT = Paediatric Autism Communication therapy. TDF = Theoretical Domains Framework. SUS = Sistema Único de Saúde (Brazilian National Public Health Service).

#### Likert ratings

The Acceptability and Appropriateness outcomes will also be assessed quantitatively using Likert ratings completed by the therapists who delivered PACT and caregivers who received PACT. These ratings were developed specifically for this study based on the conceptual domains of the IOF (Proctor et al., 2011) and are designed to capture participants’ perceptions regarding the usefulness, relevance, and overall suitability of PACT within their clinical or family contexts. The scales are presented in the Supplementary Materials.

### Other measures

Data will be collected at baseline on family sociodemographic characteristics and caregiver autistic traits and attention difficulties to characterize the sample. These data will also be used alongside other data collected during the trial, including characteristics of the SUS services and supports received as part of CAU (type of support/therapy and frequency and duration of sessions) to investigate contextual factors that affect the implementation of PACT. Sociodemographic information will be collected via an online interview with the primary caregiver to gather information on their place of residence, education level, family monthly income, whether the family receive any government benefit, working status, working hours and absenteeism rate of the adults of the family, childcare workload (time of direct care, travel time to therapies, existence of a support network), child-related expenses (cost of care and transportation), frequency of alcohol and drug consumption, race/ethnicity and the presence of physical and/or mental health problems of the caregiver and child. Autistic traits and attention difficulties in the caregiver will be assessed using the Brazilian Portuguese versions of the Short Autism Spectrum Quotient (AQ-10; Allison et al., 2012; do Nascimento Marques et al., 2025) and the Adult ADHD Self-Report Scale (ASRS, Kessler et al., 2005; Mattos et al., 2006), respectively, administered in interview format by trained researchers.

## Data management

Procedures for data management are described in the Supplementary Materials.

## Data analysis

### Analysis of effectiveness

Primary analysis will estimate the treatment effect on the primary outcome (Synchrony DCMA score) using intention-to-treat repeated-measures ANCOVA, with intervention group as the between-subjects factor and DCMA Synchrony score as the within-subjects factor repeated over time. The statistical comparison of interest in the model is the time*group interaction. Study site (i.e., the SUS service in which PACT will be delivered) will be entered as a covariate. Treatment effects and 95% confidence intervals will be reported. Missing data will be handled using maximum likelihood estimation, assuming they are missing at random. Secondary outcomes will be analysed using the same methods. Any further, secondary, analyses will be pre-specified and reviewed by the Trial Management Group.

### Sample size for effectiveness

The original PACT trial showed a treatment effect size (Cohen’s d) of 1.22 on the DCMA Synchrony measure (Green et al., 2010). A power analysis conducted in G*Power v3.1.9.7 software (Faul et al., 2007) estimated that a total sample size of 68 would be required to detect a treatment effect on the primary outcome of the same magnitude with 90% power using the planned statistical analysis, with 34 dyads allocated to PACT and 34 to Waitlist + CAU. We increased the target sample size by 50% to account for high rates of attrition that we expect from our past experience of research with PACT in Brazil (26% attrition rate in Godoy et al., 2024) as well as the complex intervention environment in which PACT will be implemented in the current trial. We therefore aim to recruit a total sample of 102 dyads, with half allocated to PACT and half to Waitlist + CAU, which allow us to detect a treatment effect size on DCMA Synchrony of 0.32 with a power of 0.90.

### Analysis of implementation

Qualitative interview data will be analysed using Reflexive Thematic Analysis following the six stages outlined by Braun and Clarke (2013). The researcher leading the implementation of PACT (PBGG) will conduct the analyses. Coding will be deductive, guided by CFIR, TDF and IOF implementation framework constructs, and inductive to allow unanticipated codes to emerge, particularly those related to sociocultural, organisational, or structural conditions of SUS that may not be captured by existing implementation frameworks. Themes will be developed iteratively from the codes with the aim of understanding, in-depth and from multiple perspectives, the contextual determinants and behavioural mechanisms of implementing PACT in SUS as well as the success of this implementation across the different regions of Brazil. Quantitative implementation outcomes will be analysed descriptively across all sites and divided by SUS service and geographical region. Exploratory analyses will examine associations between baseline caregiver characteristics and key implementation outcomes (e.g., number of sessions attended, acceptability/appropriateness ratings) to understand potential moderators of implementation trajectories.

## Discussion

This trial is the first to explore the effectiveness and implementation of an evidence-based, manualised intervention for young autistic children and their caregivers in the Brazilian public health system, at national scale. The findings from this trial will provide crucial data on the whether PACT works effectively in real-world public health clinical settings in Brazil, as well as identify factors that impede and facilitate its delivery across geographically and sociodemographically diverse SUS health services. The findings of this trial will contribute to the improvement of the care for young autistic children and their families across Brazil, potentially providing a much-needed solution to the problem of long waiting lists and inadequate or unavailable services for autism in the Brazilian public health system (dos Santos & Lima, 2024; Garcia et al., 2015; Paula et al., 2020).

Beyond the Brazilian public health context, the findings of this trial will be important for the wider scientific community working on autism interventions, particularly research focused on PACT. For example, while PACT has a robust evidence-base from large-scale randomised controlled efficacy trials (Green et al., 2010; Green, Leadbitter, Ellis et al., 2022) and small-scale clinical effectiveness trials in low-and-middle-income settings (Divan et al., 2019; Rahman et al., 2016) there has been no previous large-scale trial testing the real-world clinical effectiveness of PACT in a LMIC public health context as we propose in the current study. The findings from this trial will therefore provide critical support for proposals (Green, Leadbitter, Ainsworth et al., 2022) to adopt PACT as a primary form of post-diagnostic support for autistic children across national public health systems.

The most unique aspect of this trial, however, is that it will systematically assess the barriers and facilitators of implementing PACT in the Brazilian public health service. It is well established that demonstrating the effectiveness of an intervention is not sufficient to ensure it will be adopted in public health settings, particularly in low-and-middle-income contexts (Theobald et al., 2018). The use of elements from three implementation science frameworks to collect and analyse data from multiple perspectives on what worked in implementing PACT in SUS, and what did not work, will be crucial in designing practical implementation strategies to promote the adoption of PACT in SUS policies and services beyond the scope of this trial and in the long-term.

## Data Availability

All data produced in the present study are available upon reasonable request to the authors

## References

1. Aldred, C., Green, J., & Adams, C. (2004). A new social communication intervention for children with autism: a pilot randomised controlled treatment study suggesting effectiveness. Journal of Child Psychology and Psychiatry, 45(8), 1420–1430.

2. Aldred, C., Green, J., Howlin, P., Le Couteur, A. L., The PACT Therapists, Godoy, P.B.G.G. & Shephard, E.. (2024). Paediatric Autism Communication Therapy (PACT) Intervention Manual in Brazilian Portuguese. Hogrefe Ltd.

3. Allison, C., Auyeung, B., & Baron-Cohen, S. (2012). Toward brief “red flags” for autism screening: The short autism spectrum quotient and the short quantitative checklist in 1,000 cases and 3,000 controls. Journal of the American Academy of Child & Adolescent Psychiatry, 51(2), 202–212.

4. Benevides, T. W., Shore, S. M., Palmer, K., Duncan, P., Plank, A., Andresen, M. L., … & Coughlin, S.S. (2020). Listening to the autistic voice: Mental health priorities to guide research and practice in autism from a stakeholder-driven project. Autism, 24(4), 822–833.

5. Birken, S. A., Powell, B. J., Shea, C. M., Haines, E. R., Alexis Kirk, M., Leeman, J., … & Presseau, J. (2017). Criteria for selecting implementation science theories and frameworks: Results from an international survey. Implementation Science, 12, 124.

6. Brasil, Ministério da Saúde. (2023). Política Nacional de Atenção Integral à Saúde da Pessoa com Deficiência. Brasília: Ministério da Saúde.

7. Cane, J., O’Connor, D., & Michie, S. (2012). Validation of the theoretical domains framework for use in behaviour change and implementation research. Implementation Science, 7(1), 37.

8. Carruthers, S., Mleczko, N., Page, S., Ahuja, S., Ellis, C., Howlin, P., … & Charman, T. (2023). Using implementation science frameworks to explore barriers and facilitators for parents’ use of therapeutic strategies following a parent-mediated autism intervention. Autism, 27(4), 1011–1025.

9. Clarke, V., & Braun, V. (2013). Teaching thematic analysis: Overcoming challenges and developing strategies for effective learning. The psychologist, 26(2).

10. Curran, G. M., Bauer, M., Mittman, B., Pyne, J. M., & Stetler, C. (2012). Effectiveness-implementation hybrid designs: combining elements of clinical effectiveness and implementation research to enhance public health impact. Medical care, 50(3), 217–226.

11. Damschroder, L. J., Reardon, C. M., Opra Widerquist, M. A., & Lowery, J. (2022). Conceptualizing outcomes for use with the Consolidated Framework for Implementation Research (CFIR): The CFIR Outcomes Addendum. Implementation Science, 17, Article 7.

12. Dey, I., Chakrabarty, S., Nandi, R., Shekhar, R., Singhi, S., Nayar, S., et al. (2024). Autism community priorities in diverse low-resource settings: A country-wide scoping exercise in India. Autism, 28, 187–198.

13. Divan, G., Hamdani, S. U., Vajaratkar, V., Minhas, A., Taylor, C., Aldred, C., & Green, J. (2015). Adapting an evidence-based intervention for autism spectrum disorder for scaling up in resource-constrained settings: The development of the PASS intervention in South Asia. Global Health Action, 8(1), 27278.

14. Divan, G., Vajaratkar, V., Cardozo, P., Huzurbazar, S., Verma, M., Howarth, E., & Green, J. (2019). The feasibility and effectiveness of PASS Plus, a lay health worker delivered comprehensive intervention for autism spectrum disorders: Pilot RCT in a rural low and middle income country setting. Autism Research, 12(2), 328–339.

15. do Nascimento Marques, L., Murray, C., Fortaleza, L., Landeira Fernandez, J., & Anunciação, L. (2025). Psychometric evaluation of two adult autism screening tools in Brazil. Autism Research, 18(9), 1840–1850.

16. Faker, K., de Paula, V. A. C., & Tostes, M. A. (2024). Cross-cultural adaptation and psychometric properties of the Brazilian version of the Autism Family Experience Questionnaire (AFEQ) quality of life instrument. ABCS Health Sciences. Advance online publication. 10.7322/abcshs.2024101.28281

17. Faul, F., Erdfelder, E., Lang, A.-G., & Buchner, A. (2007). G*Power 3: A flexible statistical power analysis program for the social, behavioral, and biomedical sciences. Behavior Research Methods, 39(2), 175–191.

18. Flenik, T. M. N., Bara, T. S., & Cordeiro, M. L. (2023). Family functioning and emotional aspects of children with autism spectrum disorder in Southern Brazil. Journal of Autism and Developmental Disorders, 53, 2306–2313.

19. Fletcher-Watson, S., Adams, J., Brook, K., Charman, T., Crane, L., Cusack, J., et al. (2019). Making the future together: Shaping autism research through meaningful participation. Autism, 23(4), 943–953.

20. Geoffray, M. M., Bourgeois-Mollier, M., Maleysson-Baste, M., Gallifet, N., Dochez, S., Bonis, G., … & Jurek, L. (2025). Feasibility of a videoconferencing-based parent-mediated intervention: a mixed-method pilot study. Frontiers in Psychology, 15, 1450455.

21. Godoy, P. B. G., McWilliams, L., da Silveira, L. R., Biasão, M. C. R., Alarcão, F. S. P., Seda, L., … & Shephard, E. (2024). Acceptability and feasibility of a parent-mediated social-communication therapy for young autistic children in Brazil: A qualitative implementation study of Paediatric Autism Communication Therapy. Autism, 28(1), 123–137.

22. Godoy, P. B. G., & Shephard, E. (2025, May 28). Implementation and Effectiveness Study of a Parent-Mediated Intervention for Autistic Children in the Brazilian Health System. 10.17605/OSF.IO/EC6S4

23. Green, J., Charman, T., McConachie, H., Aldred, C., Slonims, V., Howlin, P., … & Pickles, A. (2010). Parent-mediated communication-focused treatment in children with autism (PACT): A randomized controlled trial. The Lancet, 375(9732), 2152–2160.

24. Green, J., Leadbitter, K., Ainsworth, J., & Bucci, S. (2022). An integrated early care pathway for autism. The Lancet Child & Adolescent Health, 6(5), 335–344.

25. Green, J., Leadbitter, K., Ellis, C., Taylor, L., Moore, H. L., Carruthers, S., … & Pickles, A. (2022). Combined social communication therapy at home and in education for young autistic children in England (PACT-G): a parallel, single-blind, randomised controlled trial. The Lancet Psychiatry, 9(4), 307–320.

26. Hyman, S. L., Levy, S. E., & Myers, S. M. (2020). Identification, evaluation, and management of children with autism spectrum disorder. Pediatrics, 145(1), e20193447.

27. Kessler, R. C., Adler, L., Ames, M., Demler, O., Faraone, S., Hiripi, E., et al. (2005). The World Health Organization Adult ADHD Self-Report Scale (ASRS): A short screening scale for use in the general population. Psychological Medicine, 35, 245–256.

28. LaPoint, S., Klein, C. B., Sandbank, M., Bottema Beutel, K., Fletcher Watson, S., Divan, G., … & Green, J. (2025). Maximizing the Quality and Reporting Standards of Autism Intervention Science. Autism research, 18(11), 2166–2173.

29. Leadbitter, K., Aldred, C., McConachie, H., Le Couteur, A., Kapadia, D., Charman, T., et al. (2018). The Autism Family Experience Questionnaire (AFEQ): An ecologically valid, parent-nominated measure of family experience, quality of life and prioritized outcomes for early intervention. Journal of Autism and Developmental Disorders, 48(4), 1052–1062.

30. Luyten, P., Mayes, L. C., Nijssens, L., & Fonagy, P. (2017). The parental reflective functioning questionnaire: Development and preliminary validation. PLOS ONE, 12(5), e0176218.

31. Moreira, H., & Fonseca, A. (2023). Measuring parental reflective functioning: Further validation of the Parental Reflective Functioning Questionnaire in Portuguese mothers of infants and young children. Child Psychiatry & Human Development, 54(4), 1042–1054.

32. National Institute for Health and Care Excellence. (2013). *Autism spectrum disorder in under 19s: Support and management* (NICE Guideline CG170). https://www.nice.org.uk/guidance/cg170

33. National Institute for Health and Care Excellence. (2011, updated 2021). *Autism spectrum disorder in under 19s: Recognition, referral and diagnosis* (NICE Guideline CG128). https://www.nice.org.uk/guidance/cg128

34. Pellicano, E., Dinsmore, A., & Charman, T. (2014). What should autism research focus upon? Community views and priorities from the United Kingdom. Autism, 18(7), 756–770.

35. Pickles, A., Harris, V., Green, J., Charman, T., Aldred, C., Slonims, V., … & McConachie, H. (2015). Treatment mechanism in the MRC Preschool Autism Communication Trial: Implications for study design and parent-focussed therapy for children. Journal of Child Psychology and Psychiatry, 56(2), 162–170.

36. Pickles, A., Le Couteur, A., Leadbitter, K., Salomone, E., Cole-Fletcher, R., Tobin, H., et al. (2016). Parent-mediated social communication therapy for young children with autism (PACT): Long-term follow-up of a randomized controlled trial. The Lancet, 388(10059), 2501–2509.

37. Proctor, E., Silmere, H., Raghavan, R., Hovmand, P., Aarons, G., Bunger, A., Griffey, R., & Hensley, M. (2011). Outcomes for implementation research: Conceptual distinctions, measurement challenges, and research agenda. Administration and Policy in Mental Health and Mental Health Services Research, 38(2), 65–76.

38. Rahman, A., Divan, G., Hamdani, S. U., Vajaratkar, V., Taylor, C., Leadbitter, K., &, et al. (2016). Effectiveness of the parent-mediated intervention for children with autism spectrum disorder in South Asia in India and Pakistan (PASS): A randomised controlled trial. The Lancet Psychiatry, 3(2), 128–136.

39. Rutter, M., Bailey, A., & Lord, C. (2003). The social communication questionnaire: Manual. Los Angeles, CA: Western Psychological Services.

40. Sandbank, M., Bottema-Beutel, K., Crowley, S., Cassidy, M., Dunham, K., Feldman, J. I., … & Woynaroski, T. G. (2020). Project AIM: Autism intervention meta-analysis for studies of young children. Psychological Bulletin, 146(1), 1–29.

41. Staniszewska, S., Brett, J., Simera, I., Seers, K., Mockford, C., Goodlad, S., … & Tysall, C. (2017). GRIPP2 reporting checklists: Tools to improve reporting of patient and public involvement in research. Research Involvement and Engagement, 3, 13.

42. Tafla, T. L., Rasia Lira, P., Godoy, P. B. G. & Shephard, E. (2026). Conducting meaningful participatory autism research in Latin America: an example of contextual challenges and lessons learned from the Indigenous Support Network in Brazil. Auism.

43. Theobald, S., Brandes, N., Gyapong, M., El-Saharty, S., Proctor, E., Diaz, T., … & Peters, D. H. (2018). Implementation research: new imperatives and opportunities in global health. The Lancet, 392(10160), 2214–2228.

44. World Health Organization. (2024). The World Health Organization-Five Well-Being Index (WHO-5). World Health Organization. https://creativecommons.org/licenses/by-nc-sa/3.0/igo/

