## Supplementary Material for "Trial protocol: A hybrid effectiveness-implementation study of Paediatric Autism Communication Therapy (PACT) in the Brazilian public health system"

| Description of the PACT intervention: | p.2 |
| --- | --- |
| Modifications of the PACT protocol for the Brazilian context: | p.3 |
| Safety monitoring procedures: | p.5 |
| Data management procedures: | p.11 |
| CFIR-based interview guide for service managers: | p.12 |
| Interview guide for PACT therapists: | p.14 |
| Interview guide for caregivers: | p.16 |
| Likert ratings for PACT therapists: | p.18 |
| Likert ratings for caregivers: | p.20 |

**The original PACT intervention (Green et al., 2010)**

PACT is a caregiver-mediated therapy designed to improve social-communication development in young autistic children with a wide ability range, including those with co-occurring intellectual disability and minimal language (Green et al., 2010). As a developmental intervention, PACT targets specific social-communication skills in the order in which they emerge during development. These are: 1) establishing shared attention between the caregiver and child; 2) enhancing synchrony and sensitivity in caregiver-child communication; 3) enhancing the child’s comprehension of language and communication by helping the caregiver to model language and recognise/understand the child’s communication attempts; 4) establishing the use of predictable interactive play routines to allow the child to develop anticipation of enjoyable social interactions and initiate communication in predictable situations; 5) extending the child’s communicative ability to a wider range of functions (requesting, directing) and the child’s awareness of the caregiver’s responses; 6) expanding the child’s language and communication ability to conversational turn-taking, descriptive language and narrative. Different to many other early interventions, these abilities are promoted by modifying the *caregiver’s behaviour* and the child’s sociocommunicative *environment*, rather than teaching the child to adopt different behaviours. As such, PACT is well aligned with the neurodiversity movement and with autistic community priorities for interventions.

PACT consists of 12 fortnightly sessions between the autistic child’s caregiver and a trained therapist plus 30 minutes of daily home practice. During the fortnightly sessions, the therapist supports the caregiver in learning how to increase their sensitivity to their child’s social-communication abilities, adapt their communication style to suit that of their child, and to implement strategies such as *following the child’s interests* and *watchful waiting* to support their child’s social-communication development. To facilitate the caregiver’s learning, the therapist uses video-feedback of play-based interactions between the parent and child.

The first PACT session is an initial assessment of caregiver beliefs, attitudes, therapy goals, and aspirations for the child. Subsequent sessions are structured as follows:

- Start (review of home progress, 10 minutes): caregiver feedback and discussion with the therapist about the use of strategies at home, progress, goals achieved, practice frequency, and any obstacles faced.
- Middle (video-feedback discussion, 30-60 minutes): the caregiver and therapist watch a video-recorded naturalistic interaction of the caregiver and child (pre-recorded by the caregiver or recorded during the session). Next, the therapist uses clips of the video-recording to help the caregiver identify how their social interaction and communication behaviour support the child’s social communication development and adopt the PACT strategies. The therapist and caregiver also discuss how the caregiver can practice the strategies outside of therapy sessions, in daily life routines with the child.
- End (summary and goal-setting, 10-15 minutes): the therapist summarises the main discussions of the session and helps the caregiver set goals for the home practice. How the goals (PACT strategies) will be incorporated in the caregiver’s daily interactions with the child is explored in depth, with the therapist helping to identify suitable activities and moments of the day in which to practice. The caregiver receives the home practice plan at the end of the session. The therapist also records all session details in a separate form, including the amount of home practice completed by the caregiver and the strategies they have achieved.

**Brazilian adaptation of PACT (Godoy et al., 2024)**

To implement PACT in Brazil, a study was conducted involving qualitative methods and the application of the original PACT protocol to a series of 15 autistic children aged 2 to 7 years (Godoy et al., 2024). First, the PACT manual and other materials were translated from English into Brazilian Portuguese by three bilingual (Brazilian Portuguese/English) researchers in Brazil with experience in autism and child development. Next, 18 parents of autistic children aged 2 to 10 years were recruited from child mental health clinics, and 20 clinicians (psychologists, speech therapists, occupational therapists) who work therapeutically with autistic children were recruited from national clinics offering assessment and/or treatment services for autism. Following a detailed explanation of PACT, semi-structured interviews about the intervention and its potential use in Brazil were conducted with the 18 parents and 20 clinical professionals. Concurrently, PACT was delivered to a series of cases involving 15 parent-child dyads. At the end of the PACT sessions, the 15 parents who received the intervention were also interviewed to evaluate their opinions about and experiences with PACT. All interviews were conducted individually, by telephone, audio-recorded, and subsequently analyzed according to the six stages of reflective thematic analysis by Braun & Clarke (2006).

The results showed that all parents and clinicians had favorable opinions regarding the feasibility and effectiveness of a parent-mediated intervention that can be conducted primarily at home. However, parents and clinicians also highlighted potential obstacles to the use of PACT in Brazil, particularly regarding the involvement of Brazilian parents in a caregiver-mediated therapy. Qualitative analyses revealed that parents in Brazil are often too busy with daily life tasks to act as therapists for their child and that they are more familiar with therapist-led interventions, in which a clinical professional works directly with the child. These concerns were partially confirmed in the case series of 15 families who received PACT. Based on these data, in collaboration with the UK PACT team and through discussions with researchers who implemented PACT in South Asia, minor adaptations were made to the original PACT protocol for the Brazilian context. The adaptations aimed to improve parental engagement and understanding of the intervention and included:

1) Expanded psychoeducation module delivered in the introductory PACT session to help Brazilian parents understand child communication, autism and how PACT works.
2) Altered frequency of PACT sessions to improve caregiver engagement. Instead of the 12 fortnightly sessions used in the original PACT protocol, the Brazilian protocol delivers the first four sessions weekly and then the remaining eight sessions fortnightly.
3) Culturally adapted real-life examples of PACT strategies and suggestions of toys for caregiver-child interactions, relevant to the Brazilian context.
4) Increased emphasis on the importance of the therapist helping the caregiver address challenges in findings a suitable time and place in the home to participate in PACT.
5) Audio-recorded practice programme for families who cannot read.

**Protocol for monitoring adverse events *(translated to English for publication)***

This document described the standardised procedures for identifying, recording, monitoring and reporting adverse events and serious adverse events in the study investigating the implementation and effectiveness of Paediatric Autism Communication Therapy (PACT) in the Brazilian public health system (SUS).

PACT is a low-risk intervention, but the study will adopt active and passive surveillance to identify and respond rapidly to adverse events during the research.

**1. Definitions**

Adverse event: any unwanted/undesirable medical, psychological or social event that happens to the participant during the study, not necessarily related to the intervention.
Serious adverse event: an event that results in death, risk to life, hospitalisation or serious disability, or a serious event related to child protection.
Relation to the intervention: not related / probably not related / possibly related / probably related / definitely related.

**2. Adverse events of special interest to this study**

• Increase of stress or distress in the caregiver or child
• Family conflict associated with changes in routine
• Suicidal thinking in the caregiver
• Child protection incident
• Data violation
• Interference related to treatment as usual

**3. Detection protocol**

Active surveillance: checklist of adverse events completed by the therapist at the end of each session.
Passive surveillance: spontaneous reporting.

**4. Documentation**

All adverse events and serious adverse events will be recorded on a form, including a description of the event, date, level of seriousness, relation to the intervention, and conduct.

**5. Reporting deadlines**

• Serious adverse events, data violations or child protection issues will be notified within 24h to the principal investigators (Priscilla Godoy or Elizabeth Shephard) and within 72h to the Ethics Committee.

• Non-serious adverse events will be registered on a form within 7 days.

**6. Safety governance**

The investigator Priscilla Godoy and the Data Safety and Monitoring Board will revise all adverse event forms completed by therapists. The Data Safety and Monitoring Board will also review a summary of adverse event data (compiled by Priscilla Godoy) every 4 months.

**7. Integration with the implementation study of PACT**

Barriers and difficulties related to the implementation of PACT will also be monitored, including therapist overburden, logistical failures and deviations from the PACT protocol.

**8. Summary of the flow of adverse event procedures**

1. Detection of an adverse event (during a PACT session, telephone contact or by spontaneous report).
2. Immediate screening and forwarding for clinical attention if necessary.
3. Recording on adverse event record form (within 24h for serious adverse event, 7 days for non-serious adverse event).
4. Review by the principal investigator and the Data Safety and Monitoring Board, and final classification of the event.
5. Inclusion in 4-monthly report.

**Adverse event checklist for completion at the end of each PACT session *(translated to English for publication)***

Complete this form at the end of each PACT session. In case of any adverse event, record the details on the Adverse Event Form e follow the instructions on the protocol for adverse events.

**Identification**

Participant ID: ________________________________

Session date: ____/____/______     Session number: _______

Therapist name: __________________________________________

1. Did the participant (child) present emotional or behavioural discomfort during the session?  (  ) No   (  ) Yes. If yes, describe:_______________________________________________________________

2. Did the caregiver present significant anxiety, stress or frustration during or after the session?  (  ) No  (  ) Yes. If yes, describe:_______________________________________________________________

3. Were there family conflicts or misunderstandings or important interruptions in family life?  (  ) No  (  ) Yes. If yes, describe:_______________________________________________________________

4. Was there a spontaneous report of clinical decrement, change in medication or other recent medical event?  (  ) No  (  ) Yes. If yes, describe:_______________________________________________________________

5. Was there any technical difficulty, confidentiality leak or problem with video recording?   (  ) No  (  ) Yes. If yes, describe:_______________________________________________________________

6. Any other relevant event?  (  ) No  (  ) Yes. If yes, describe:______________________________________________________________

**Other observations (if needed)**

______________________________________________________________________
______________________________________________________________________
______________________________________________________________________

**Next steps**

☐ None  ☐ Telephone follow up  ☐ Forward for clinical treatment  ☐ Other: __________________________

**Adverse Event / Serious Adverse Event Record Form *(translated to English for publication)***

Complete this form for every occurrence of an adverse event or serious adverse event.

**1. Participant identification**

Participant ID: ________________________________

Age: ______  Sex: ______  Region/Service name: ____________________________

**2. Event information**

Start date: ____/____/______     End date: ____/____/______

Detailed description of the event: ____________________________________________________________________________________________________________________________________________
______________________________________________________________________

**3. Classification**

(  ) Non-serious  (  )  Moderate  (  )  Serious

(  )   Expected  (  )   Unexpected

Relation to the intervention: (  )   Not related (  )   Probably unrelated  (  )   Possibly related  (  )   Probably related (  )   Definitely related

**4. Conduct and outcome**

Actions taken: _______________________________________________

Forwarding: _____________________________________

Outcome: (  )   Resolved  (  )   In resolution  (  )   Permanent consequences  (  )   Death

Date of resolution: ____/____/______

**5. Notification and review**

Detected by: (  ) Therapist    (  ) Caregiver    (  ) Other (specify): _____________

Date of initial notification: ____/____/______   Time: ________

Assessment by the principal investigator: _________________  Date: ____/____/______

Assessment by the Data Safety and Monitoring Board: _______________ Date: ____/____/______

Signature: ____________________________

**Data Management**

Personal information, including names, addresses, contact information by email and telephone, date of birth, and sex, will be collected about the children and their primary caregivers. This information will be retained only after primary caregivers provide written informed consent and sign the consent form to enter the study. Information will be stored in an encrypted electronic database on the secure online platform, Google Workspace, of the University of São Paulo, with access granted only to members of the research team or other authorized personnel (e.g., auditors). Video recordings of the caregiver-child and adult-child interaction assessment and PACT intervention sessions will be saved only with the participants' anonymous identification number and will be stored electronically in an encrypted storage system on the secure Google Workspace platform of the University of São Paulo, with access granted only to members of the research team or other authorized personnel. All data generated by the study will be preserved in secure storage at the Institute of Psychology, University of São Paulo, for at least five years after the project's completion. Anonymous data files will be made available for sharing with other research groups in Brazil and internationally, provided that (a) families agree that their data will be shared in this way during the consent process, and (b) data are shared securely via encrypted electronic data sharing sites, specifically the Google Workspace of the University of São Paulo.

**Interview guide for SUS service managers / coordinators based on the Consolidated Framework for Implementation Research (CFIR) *(translated to English for publication)***

**Aim**: To understand organisational and strategic barriers and facilitators related to the implementation of PACT within the service.

**Estimated Duration**: 30–40 minutes.

**Introduction**
We would like to talk about how your service is preparing to implement PACT and which factors may facilitate or hinder this process. There are no right or wrong answers. Our goal is to understand how the service operates and to hear your perspectives.

**Question Blocks**

| **CFIR Domain** | **Suggested Questions** |
| --- | --- |
| 1. Intervention Characteristics | “What is your understanding of PACT and its key components?”  “Do you believe this intervention is well suited to the families served here?”  “How easy or difficult do you think it will be to integrate PACT into the service’s routine?” |
| 2. Outer Setting | “What are the main demands you receive from families of autistic children?”  “Are there any incentives or requirements from municipal or state authorities to offer this type of intervention?”  “How does the service coordinate with other care networks (e.g., schools, CAPS-IJ, NASF?”  “Does the service currently have a waiting list or increased demand for autistic children requiring care?” |
| 3. Inner Setting | “How would you describe the work culture here? Does the team generally welcome new practices?”  “Is there adequate physical space for video recording?”  “Would the length of your standard sessions allow for incorporating this approach?”  “How would you describe the current team climate?” |
| 4. Characteristics of Individuals | “How are the professionals feeling about the introduction of PACT?”  “Do they typically engage in training and capacity-building activities?”  “Is there anyone who seems more resistant or may require additional support?” |
| 5. Process (planning) | “How does the service plan to integrate PACT into its routine?”  “We will offer monthly 1-hour group supervision sessions for therapists - how do you think this would work within the daily realities of the service?”  “Will the service adopt any strategies to monitor the delivery or progress of the intervention?” |

**Closing**

“Is there anything else you consider important for ensuring that PACT is implemented effectively in your service?”

**Interview guide for SUS therapists concerning PACT delivery *(translated to English for publication)***

**Theoretical Basis**: Theoretical Domains Framework (TDF) + Implementation Outcomes Framework (IOF).

**Aim**: To understand therapists’ perceptions regarding the implementation process, barriers and facilitators, and perceived outcomes (acceptability, appropriateness, feasibility, fidelity, and sustainability).

**Estimated Duration**: 45–60 minutes.

**Introduction**

We would like to talk about your experience delivering PACT to the families in this service. Our aim is to understand what worked well, what was challenging, and how you perceive the effects of this intervention within the context of the Brazilian public health system (SUS). There are no right or wrong answers - we are interested in hearing about your genuine, personal experience.

**Question Blocks**

| **Theoretical Axis** | **Theme / Domain** | **Suggested Questions** |
| --- | --- | --- |
| A. General experience with PACT (IOF – Acceptability, Appropriateness) | Initial and overall perceptions of the intervention | “Overall, how was your experience delivering PACT?” / “What did you find most useful or relevant in the intervention?” / “Do you think PACT is suitable for the type of families you work with?” |
| B. Practical conditions (IOF – Feasibility) | Resources, time, and routine | “Was it feasible to deliver PACT within the conditions of your service?” / “How was it to balance the sessions with your other work demands?” / “What helped or made it more difficult?” / “Are there aspects of the service that make it harder to carry out this type of intervention, which requires a more relational approach?” / “How do you feel in your work when delivering this type of intervention?” |
| C. Fidelity and adaptation (IOF – Fidelity / Adaptability) | Adherence to the protocol | “Were you able to follow the steps as outlined in the manual?” / “Did you make any adaptations to fit the realities of the families or the service?” / “How did you decide what to adapt?” |
| D. Perceived outcomes and impact (IOF – Adoption / Penetration / Sustainability) | Effects on professionals, families, and services | “What changes did you observe in the families you worked with using PACT?” / “Did you notice any impact on team dynamics or ways of working?” / “Do you intend to continue using PACT? Why?” |
| E. Individual mechanisms (TDF – Knowledge, Skills, Beliefs) | Changes in practice and learning | “What did you learn from this experience?” / “In what ways did PACT influence how you view caregiver–child interaction?” / “Do you feel more confident in guiding caregivers?” |
| F. Emotions and attitudes (TDF – Emotion, Optimism) | Feelings associated with the process | “How did you feel during the sessions?” / “Were there any particularly rewarding or frustrating moments?” / “What motivated you to continue?” |
| G. Social influences and support (TDF – Social Influence / Context) | Team climate and institutional support | “How did your colleagues and managers respond to PACT?” / “Was there sufficient support from leadership?” / “Do you think the service is prepared to sustain the intervention?” |
| H. Sustainability (IOF – Sustainability) | Continuity and integration | “Do you think PACT can be sustained as part of routine practice within the service?” / “What would be needed to ensure this?” |
| I. Suggestions and final reflections | Recommendations for improvement | “What would you change in the implementation process or in the training?” / “Is there anything that should be done differently to make it easier for professionals to deliver the intervention?” |

**Closing**

“Is there anything else you would like to share about your experience with PACT, or anything you consider important for ensuring its effective implementation in public health services?”

**Interview guide for caregivers about their experiences of receiving PACT *(translated to English for publication)***

**Theoretical basis**: Theoretical Domains Framework (TDF) + Implementation Outcomes Framework (IOF).
**Aim**: To understand caregivers’ perceptions regarding the process, engagement, perceived effects on the caregiver–child relationship, and the feasibility of implementing the intervention at home.

**Estimated duration**: 40–50 minutes.

**Introduction**

We would like to hear about your experience receiving PACT - what you thought about the sessions, what may have changed in your daily routine with your child, and how it was to put what you learned into practice. Everything you share is confidential and valuable for improving the program.

| **Theoretical Axis** | **Theme / Domain** | **Suggested Questions** |
| --- | --- | --- |
| A. General experience (IOF – Acceptability) | Perception of the intervention | “How was it for you to take part in PACT?” / “What did you like most, or find most difficult, about the sessions?” |
| B. Clarity and fit (IOF – Appropriateness) | Meaning and relevance | “Do you feel that PACT matched your family’s needs?” / “Did the content make sense to you and your child?” |
| C. Everyday applicability (IOF – Feasibility) | Feasibility of use at home | “Was it easy or difficult to put into practice what you learned in the sessions?” / “What helped or made it harder within your daily routine?” |
| D. Adoption and continuity (IOF – Adoption / Sustainability) | Maintenance of strategies | “Did you continue using the strategies after the sessions ended?” / “Do you plan to continue?” / “Why?” |
| E. Perceived effects (IOF – Perceived consequences) | Changes in communication | “What differences have you noticed in communication with your child since starting PACT?” / “Did anything surprise you?” |
|  | Changes in the relationship | “Do you feel that your relationship with your child has changed over the course of the intervention? How?” / “Is there anything you do now with your child that you didn’t do before?” / “When your child does something you don’t understand or find difficult, what do you usually do now?” |
|  | Responsiveness and attunement | “Do you feel that you can better understand what your child wants or is trying to communicate?” / “What do you do when your child tries to communicate with you?” / “Do you feel that you need to direct or control interactions less now?” / “What do you do when the interaction doesn’t ‘flow’ or becomes difficult to continue?” |
|  | Secure base | “When your child feels more insecure or upset, what usually happens between you?” / “How does your child respond when they need help or support?” / “Has this changed since the intervention?” |
|  | Predictability of the relationship | “Do you feel that you can better anticipate your child’s reactions now?” / “Do your interactions feel easier, more difficult, or different than before?” |
|  | Meaning of the child’s behaviour | “When your child does something difficult or challenging, how do you understand it now?” / “Do you feel that the way you interpret your child’s behaviour has changed?” |
|  | Caregiver emotional experience | “How do you feel during interactions with your child now?” / “Compared to the beginning, what has changed for you emotionally?” |
| F. Self-efficacy and skills (TDF – Beliefs about capabilities / Skills) | Confidence in applying strategies | “Do you feel able to use what you learned on your own now?” / “Is there anything that still feels difficult?” |
| G. Emotions and motivation (TDF – Emotion / Reinforcement) | Affective responses and motivation | “How did you feel during the sessions?” / “What kept you motivated to continue?” / “Was there any moment that stood out to you?” |
| H. Family context (TDF – Environmental context / Social influences) | Support and external barriers | “Did anyone in your family support you in taking part?” / “Was there anyone who didn’t understand or support the process?” |
| I. Suggestions (improvement and expansion) | Overall evaluation | “What would you change about how PACT was delivered?” / “Would you recommend this intervention to other families?” / “Why?” |

**Closing**

Thank you for sharing your experience. What you have told us will help us understand what works best for families and how we can improve PACT for others.

**Likert rating for SUS therapists to assess the appropriateness and acceptability of PACT *(translated to English for publication)***

Please read each statement about your experience with the PACT intervention carefully and indicate to what extent you agree with each one. There are no right or wrong answers - what is important to us is your personal perception.

Use the following scale to mark your response:

1 = Completely disagree   2 = Disagree   3 = Neither agree nor disagree   4 = Agree   5 = Completely agree

Mark only one option per line, choosing the number that best represents your opinion. Your responses are confidential and will be used only for research, helping us to understand how the intervention works in different contexts and for different people.

| Item | Completely disagree | Disagree | Neither agree nor disagree | Agree | Completely agree |
| --- | --- | --- | --- | --- | --- |
| 1. In general, I consider PACT acceptable for use in my clinical practice. |  |  |  |  |  |
| 2. I like the format of PACT sessions (video-feedback, focus on the caregiver-child interaction). |  |  |  |  |  |
| 3. I am satisfied with the way in which I was trained to deliver PACT. |  |  |  |  |  |
| 4. I feel comfortable delivering PACT to the families in my service. |  |  |  |  |  |
| 5. I would recommend PACT to other professionals of my service. |  |  |  |  |  |
| 6. PACT is appropriate for the needs of the autistic children and families I work with. |  |  |  |  |  |
| 7. PACT is compatible with the objectives and routine of my service. |  |  |  |  |  |
| 8. PACT is aligned with my way of working with autistic children and their families. |  |  |  |  |  |
| 9. Using PACT makes sense considering the resources available at my service. |  |  |  |  |  |
| 10. PACT appears to be an appropriate intervention for the Brazilian public health system (SUS). |  |  |  |  |  |

**Likert rating for caregivers to assess the appropriateness and acceptability of PACT *(translated to English for publication)***

Please read each statement about your experience with the PACT intervention carefully and indicate to what extent you agree with each one. There are no right or wrong answers - what is important to us is your personal perception.

Use the following scale to mark your response:

1 = Completely disagree   2 = Disagree   3 = Neither agree nor disagree   4 = Agree   5 = Completely agree

Mark only one option per line, choosing the number that best represents your opinion. Your responses are confidential and will be used only for research, helping us to understand how the intervention works in different contexts and for different people.

| Item | Completely disagree | Disagree | Neither agree nor disagree | Agree | Completely agree |
| --- | --- | --- | --- | --- | --- |
| 1. In general, I am satisfied with the PACT intervention. |  |  |  |  |  |
| 2. I liked the way the PACT sessions were conducted (watch the videos and discuss with the therapist). |  |  |  |  |  |
| 3. I felt comfortable participating in the PACT sessions. |  |  |  |  |  |
| 4. I feel that I was respected and listened to during PACT. |  |  |  |  |  |
| 5. I would recommend PACT to other families of autistic children. |  |  |  |  |  |
| 6. PACT was appropriate for the needs of my child. |  |  |  |  |  |
| 7. PACT fitted in well with our family’s daily routines. |  |  |  |  |  |
| 8. The strategies suggested in PACT made sense to me. |  |  |  |  |  |
| 9. What I learned in PACT is consistent with how I would like to relate to my child. |  |  |  |  |  |
| 10. PACT was the right type of support for our family at this time. |  |  |  |  |  |
